# Nodules as a Risk Factor for Acute Attacks in Lymphedema: Evidence from Addis Ababa City Administration, Ethiopia

**DOI:** 10.1101/2025.05.12.25327479

**Authors:** Mebratu Mitiku Jemberie, Tsige Amberbir Wondimagegnehu, Abebe kelemework Takele, Mulat Tilahun Belay, Fikre Hailekiros Gidey, Fetiya Kedir Abamegal, Merga Mekonnen Akuma, Gail Davey

## Abstract

**Background:** Lymphedema is a chronic condition characterized by fluid accumulation and tissue swelling, often complicated by recurrent acute attacks. Nodules, a common feature of chronic lymphedema, may contribute to the frequency and severity of these episodes. However, nodules as a risk factor for acute attacks remains poorly understood. This study aimed to assess the prevalence of nodules, the frequency of acute attacks, and nodule as risk factor for acute attacks among individuals with lymphedema in Addis Ababa city Administration, Ethiopia.

**Methods:** A cross-sectional study was conducted among individuals with lymphedema in Addis Ababa city Administration. Data were collected using structured interviews and clinical examinations. The prevalence of nodules and acute attacks was determined, and logistic regression was used to assess their association.

**Results:** Among the participants, 57.5% reported the presence of nodules affecting multiple areas of the lower limbs. Additionally, 41.2% experienced acute attacks, with an average of 5.89 episodes in the past six months. Logistic regression analysis revealed that individuals with nodules were 3.2 times more likely to experience acute attacks compared to those without nodules (p = 0.001). Other demographic and clinical factors, including sex and lymphedema severity, also showed trends associated with acute attack occurrence. Stigma was a significant concern, with 42.6% of participants reporting stigma from their communities (60.9%) and families (36.8%). Additionally, 34.8% of participants faced workplace challenges due to their condition.

**Conclusions:** The occurrence of nodules was associated with an increased risk of acute attacks. Both nodules and acute attacks were commonly observed. Many patients faced stigma from close family members and the surrounding community.

**Author Summary:** Lymphedema is a long-term condition that leads to swelling in the limbs due to fluid accumulation and impaired lymphatic function. People with lymphedema often experience recurrent acute attacks, which cause pain, inflammation, and disability. This study investigated the relationship between nodules—hardened tissue formations commonly found in chronic lymphedema—and the occurrence of acute attacks among individuals with lymphedema in Addis Ababa City Administration, Ethiopia.

Our findings revealed that more than half of the participants had nodules, and over 40% experienced acute attacks, with an average of nearly six episodes in the past six months. We found that individuals with nodules were significantly more likely to suffer from acute attacks, suggesting that nodules may serve as reservoirs for bacteria or contribute to lymphatic obstruction. Additionally, many participants reported experiencing stigma from their communities and families, as well as difficulties in the workplace due to their condition.

These findings highlight the need for early detection and management of nodules to prevent acute attacks, improve patient quality of life, and reduce the psychosocial and economic burden of lymphedema. Public health interventions should focus on improving self-care practices, providing accessible treatment options, and addressing stigma through education and community awareness programs. Future research should explore long-term strategies for preventing and managing nodules to reduce complications associated with lymphedema.

## Introduction

Lymphedema is a chronic and debilitating condition caused by impaired lymphatic drainage, leading to progressive swelling, fibrosis, and recurrent inflammatory episodes in the affected limb(s) [1, 2]. Globally, lymphedema is most commonly associated with neglected tropical diseases (NTDs) such as lymphatic filariasis (LF) and podoconiosis, both of which are endemic in Ethiopia [3, 4]. LF is caused by filarial parasites transmitted through mosquito bites, whereas podoconiosis results from long-term exposure to red clay soil with high mineral content, triggering an inflammatory response in genetically susceptible individuals [5, 6]. These conditions contribute significantly to morbidity and disability in affected populations, often leading to reduced mobility, psychosocial distress, and economic hardship [7, 8].

Among individuals with lymphedema, acute dermatolymphangioadenitis (ADLA), commonly referred to as acute attacks, represents one of the most frequent and debilitating complications [9]. Acute attacks are characterized by sudden episodes of fever, pain, erythema, and swelling, often leading to functional impairment, hospitalization, and further deterioration of lymphatic function [10, 11]. Studies suggest that bacterial infections, poor skin hygiene, and chronic lymphatic inflammation are key contributors to acute attacks [12, 13]. Consequently, effective morbidity management, including hygiene promotion, wound care, and antibiotic therapy, is critical in reducing their frequency and severity [14, 15].

One of the underexplored clinical features of lymphedema is the presence of nodules—firm, localized tissue formations that may develop due to chronic inflammation, fibrosis, and recurrent infections [16]. Nodules have been observed in advanced-stage lymphedema, particularly in individuals with long-standing disease progression [17, 18]. Despite their frequent occurrence, the relationship between nodules and acute attacks remains poorly understood. Several hypotheses suggest that nodules may serve as bacterial reservoirs, exacerbate lymphatic obstruction, or trigger persistent inflammatory responses, increasing susceptibility to recurrent acute attacks [19, 20]. However, limited research has quantitatively assessed this association, particularly in low-resource settings such as Ethiopia [21].

Ethiopia bears a significant burden of LF- and podoconiosis-related lymphedema, with an estimated one million affected individuals [22]. Despite ongoing morbidity management and disability prevention (MMDP) programs, gaps remain in the early detection, treatment, and long-term care of lymphedema patients [23]. Moreover, stigma, social exclusion, and economic challenges further hinder healthcare access and treatment adherence [24, 25]. Addressing these challenges requires a deeper understanding of disease progression, risk factors for acute attacks, and effective intervention strategies [26, 27].

This research aimed to examine the connection between nodules and acute attacks in individuals with lymphedema in Addis Ababa City Administration, Ethiopia. The study focused on identifying the prevalence of nodules among those affected, assessing the frequency of acute attacks along with potential risk factors, and exploring the relationship between nodules and the occurrence of acute attacks.

By identifying key predictors of acute attacks, this study provides critical insights for improving morbidity management and informing public health interventions aimed at reducing the burden of lymphedema and improving patient outcomes. The findings contribute to evidence-based strategies for reducing the impact of LF- and podoconiosis-related disability, thereby enhancing the quality of life for affected individuals in Ethiopia and similar settings.

## Materials and Methods

### Study Design & Setting

This study employed a cross-sectional, church-, mosque-, and household-based design to assess the prevalence of acute attack, nodules, and association between them in Addis Ababa City Administration, Ethiopia. The primary objective was to examine the relationship between nodules and acute attacks. A cross-sectional study design was selected as it allows for associations between variables to be examined at a single point in time. Trained data collectors from local churches and mosques facilitated comprehensive case identification to ensure accuracy and inclusivity. The study was conducted across Addis Ababa’s 11 sub-cities, covering 265 churches, 352 mosques, and 32 households. The selection of churches and mosques as data collection sites was based on the congregation of individuals affected by lymphedema and hydrocele, many of whom migrate from rural areas and seek shelter in religious settings due to stigma and financial hardship. For those not found in these religious locations, additional tracing was conducted at the household level to ensure comprehensive case capture.

### Study Population & Sampling

The study population comprised individuals residing around churches and mosques in Addis Ababa City Administration who were suspected of having lymphedema or hydrocele. Participants were eligible regardless of age or gender if they presented with lymphedema and provided informed consent. Individuals with other forms of leg swelling unrelated to LF or podoconiosis, and those unwilling to participate were excluded from the study. A complete census approach was employed instead of random sampling, ensuring the identification and registration of all lymphedema cases within the selected churches and mosques. Data collection was facilitated by trained Sunday school students and their teachers, as well as mosque leaders who assisted in community engagement and identification of suspected cases. The Addis Ababa City Administration Health Bureau provided supervisors and health officers who verified and cross-checked the collected data for quality assurance. Individuals diagnosed with lymphedema underwent further assessment to determine the presence of nodules and their history of acute attacks.

### Data Collection

Structured questionnaires were used to collect data, encompassing demographic information such as age, sex, previous residence, marital status, and education level. Clinical characteristics, including the presence and duration of lymphedema swelling, its severity—classified as mild (swelling without skin folds and potentially reversible at night), moderate (swelling with shallow folds), or severe (swelling with skin changes such as mossy lesions, knobs, and/or deep folds)— as well as the location of the swelling, history of acute attacks within the past six months, and the presence of nodules, were recorded. Psychosocial and economic impacts were assessed through structured questions. These included a direct question to determine whether participants experienced stigma or discrimination related to their leg swelling, and questions evaluating the impact of the condition on their livelihood and daily activities. Participants were also asked to self-report their economic status by categorizing themselves as poor, medium, or rich. To ensure accurate data collection, one-day training was provided to Sunday school students, church supervisors, Ethiopian Muslims Development Agency (EMDA) members. Health officers from the Addis Ababa City Administration Health Bureau were also trained in the differential diagnosis of lymphedema, survey administration techniques, and patient counseling. Quality control measures were implemented throughout data collection. Supervisors reviewed data collection forms daily, and a random verification of 10% of recorded cases was conducted by health professionals to confirm diagnoses. Photographic documentation and visual guides were used to differentiate stages of lymphedema.

### Data Analysis

Data analysis was conducted using SPSS (Statistical Package for Social Sciences) Version 26. Descriptive statistics were applied to determine the prevalence of lymphedema, demographic distributions, and frequency of acute attacks and nodules. Inferential statistics, including chi-squared tests, were used to assess the association between nodules and acute attacks, while logistic regression analysis identified predictors of acute attacks among individuals with lymphedema.

### Ethical Considerations

Ethical considerations were strictly followed: ethical approval was obtained from the Addis Ababa City Administration Health Bureau Research Ethics Committee and participants completed informed consent forms. The study adhered to World Health Organization ethical guidelines, ensuring participant confidentiality and the right to withdraw at any time.

## Results

### Descriptive Statistics

A total of 204 participants with lower leg lymphedema volunteered to take part in the study. Data were collected from 11 sub-cities in Addis Ababa City Administration, Ethiopia. As shown in Table 1, the majority of screening activities were conducted in churches, accounting for 67.2% (137 out of 204), followed by mosques (17.2%, 35) and households (15.7%, 32).

**Table 1:** Screening areas of the study.

| Screening Area | Frequency | Percent | Valid Percent | Cumulative Percent |
| --- | --- | --- | --- | --- |
| Church | 137 | 67.2 | 67.2 | 67.2 |
| Mosque | 35 | 17.2 | 17.2 | 84.3 |
| Household | 32 | 15.7 | 15.7 | 100.0 |
| **Total** | **204** | **100.0** | **100.0** | **100.0** |

As shown in Table 2, the largest group of participants were found in Addis Ketema (27.5%, 56), followed by Kolfe Keraniyo (18.6%, 38) and Gulele (12.3%, 25). Together, Addis Ketema and Kolfe Keraniyo accounted for nearly half (46.1%) of all participants, while sub-cities like Akaki Kality and Arada had the smallest representation.

**Table 2:** Distribution of participants across sub-cities of Addis Ababa.

| Sub-City | Frequency | Percent | Cumulative Percent |
| --- | --- | --- | --- |
| Arada | 4 | 2.0 | 2.0 |
| Addis Ketema | 56 | 27.5 | 29.4 |
| Akaki Kality | 3 | 1.5 | 30.9 |
| Bole | 17 | 8.3 | 39.2 |
| Gulele | 25 | 12.3 | 51.5 |
| Kirkos | 15 | 7.4 | 58.8 |
| Kolfe Keraniyo | 38 | 18.6 | 77.5 |
| Lideta | 18 | 8.8 | 86.3 |
| Nifas Silk Lafto | 20 | 9.8 | 96.1 |
| Yeka | 8 | 3.9 | 100.0 |
| **Total** | **204** | **100.0** | **100.0** |

The study included 107 females (52.5%) and 97 males (47.5%). The participants’ ages ranged from 18 to 90 years, with a mean age of 48.8 years (SD = 14.56) as shown in Table 3. The majority participants were originated from the Amhara region (63.5%, 129), followed by South Ethiopia Region (11.3%, 23) and Addis Ababa City Administration (9.3%, 19). In terms of occupation, 51.5% (105) were beggars, followed by housewives (12.7%, 26) and daily laborers (9.8%, 20). The majority (61.3%, 125) were illiterate, and only 1.0% (2) had attained a college or university education. In terms of marital status, 47.8% (97) were married, and 20.2% (41) were single. The majority (85.8%, 175) study participants categorized them-selves as poor..

**Table 3:** Demographic Characteristics of the participants.

| **Variable** | **Category** | **Frequency** | **Percentage (%)** |
| --- | --- | --- | --- |
| **Sex** | Male | 97 | 47.5 |
|  | Female | 107 | 52.5 |
| **Age (Years)** | Range: 18–90 | - | - |
|  | Mean: 48.8 | - | - |
|  | Standard Deviation: 14.56 | - | - |
| **Region of Origin** | Amhara | 129 | 63.5 |
|  | South Ethiopia | 23 | 11.3 |
|  | Addis Ababa | 19 | 9.4 |
|  | Oromia | 17 | 8.4 |
|  | Tigray | 4 | 2.0 |

| Variable | Category | Frequency | Percentage (%) |
| --- | --- | --- | --- |
|  | Other Regions | 11 | 5.4 |
| **Occupation** | Beggar | 105 | 51.5 |
|  | Housewife | 26 | 12.7 |
|  | Daily Laborer | 20 | 9.8 |
|  | Agriculture | 15 | 7.4 |
|  | Other | 12 | 5.9 |
|  | Job-Seeker | 9 | 4.4 |
|  | Merchant | 8 | 3.9 |
|  | Private Worker | 7 | 3.4 |
|  | Student | 2 | 1.0 |
| **Education** | Illiterate | 125 | 61.3 |
|  | Write/Read (No Formal) | 30 | 14.7 |
|  | Grade 1–4 | 22 | 10.8 |
|  | Grade 5–8 | 15 | 7.4 |
|  | Grade 9–10 | 10 | 4.9 |
|  | College/University | 2 | 1.0 |
| **Marital Status** | Married | 97 | 47.8 |
|  | Single | 41 | 20.2 |
|  | Separated | 21 | 10.3 |
|  | Divorced | 19 | 9.4 |
|  | Widowed | 25 | 12.3 |
| **Wealth** | Poor | 175 | 85.8 |
|  | Medium | 29 | 14.2 |

### Clinical Characteristics

Most participants reported swelling in both legs (59.5%, 119), followed by the right leg only (19.5%, 39) and left leg only (16.5%, 33). Nodules were present in 57.5% (115) of participants, commonly affecting multiple areas (62.3%, 71). Hydrocele was reported in 4.9% (10) of participants, with an average duration of 8.77 years (SD = 11.34) as shown in Table 4. Acute attacks were experienced by 41.2% (84) of participants, with an average of 5.89 attacks (SD = 5.58) in the past six months. Regarding lymphedema severity, 42.5% (85) had moderate lymphedema, 35.5% (71) had mild cases, and 22.0% (44) had severe cases.

**Table 4:** Clinical characteristics of participants.

| **Variable** | **Category** | **Frequency** | **Percentage (%)** |
| --- | --- | --- | --- |
| **Swelling Area** | Both Legs | 119 | 59.5 |
|  | Right Leg only | 39 | 19.5 |
|  | Left Leg only | 33 | 16.5 |
|  | Left Hand | 6 | 3.0 |
|  | Right Breast | 2 | 1.0 |
|  | Right Hand | 1 | 0.5 |
| **Nodule Presence** | Yes | 115 | 57.5 |
|  | No | 85 | 42.5 |
| **Area of Nodule** | Many Areas | 71 | 62.3 |
|  | Foot Toes | 17 | 14.9 |
|  | Foot Arch | 15 | 13.2 |
|  | Heel | 11 | 9.6 |
| **Hydrocele** | Yes | 10 | 4.9 |
|  | No | 194 | 95.1 |

| Variable | Category | Frequency | Percentage (%) |
| --- | --- | --- | --- |
| **Years of Hydrocele** | Range: 0.60–30.00 years | - | - |
|  | Mean: 8.77 years | - | - |
|  | Standard Deviation: 11.34 | - | - |
| **Hydrocele Treatment** | Yes | 4 | 66.7 |
|  | No | 2 | 33.3 |
| **Acute Attacks in the last six months** | Yes | 84 | 41.2 |
|  | No | 120 | 58.8 |
| **Number of Acute Attacks in the last six months** | Range: 1–20 | - | - |
|  | Mean: 5.89 | - | - |
|  | Standard Deviation: 5.58 | - | - |
| **Lymphedema Stage** | Mild | 71 | 35.5 |
|  | Moderate | 85 | 42.5 |
|  | Severe | 44 | 22.0 |
| **Sought treatment for lymphedema in the past 2 years** | Yes | 155 | 77.5 |
|  | No | 45 | 22.5 |
| **Reasons for Not Seeking Treatment** | Lymphedema has no treatment | 10 | 23.8 |
|  | Don't know service provider | 6 | 14.3 |
|  | Treatment is costly | 11 | 26.2 |
|  | Facility is too far | 1 | 2.4 |
|  | No reason | 13 | 31.0 |
|  | Other | 1 | 2.4 |

### Inferential Statistics

#### Association between Nodules and Acute Attacks

Logistic regression analysis identified the presence of nodules as a significant predictor of acute attacks, with individuals having nodules being 3.2 times more likely to experience acute attacks compared to those without (p = 0.001). Similarly, a chi-squared test demonstrated a strong association between nodules and the frequency of acute attacks (χ² = 12.43, p < 0.001), indicating that participants with nodules were at a higher risk. Furthermore, logistic regression analysis revealed that individuals with moderate to severe lymphedema had significantly greater odds of experiencing acute attacks compared to those with mild lymphedema (OR = 3.12, 95% CI: 1.85–5.24, p < 0.001), as presented in Table 5.

**Table 5:** Inferential statistics of variables.

| Variable | B | S.E. | Wald | df | Sig. | Exp(B) |
| --- | --- | --- | --- | --- | --- | --- |
| Nodule | 1.170 | 0.349 | 11.240 | 1 | 0.001 | 3.223 |
| Sex | -0.566 | 0.338 | 2.798 | 1 | 0.094 | 0.568 |
| Age | -0.006 | 0.013 | 0.227 | 1 | 0.634 | 0.994 |
| Occupation | 0.054 | 0.070 | 0.599 | 1 | 0.439 | 1.056 |
| Education | -0.176 | 0.146 | 1.457 | 1 | 0.227 | 0.839 |
| Married | 0.116 | 0.138 | 0.707 | 1 | 0.400 | 1.123 |
| Wealth | 0.488 | 0.501 | 0.947 | 1 | 0.330 | 1.628 |
| Years of Swelling | 0.019 | 0.015 | 1.550 | 1 | 0.213 | 1.019 |
| Lymphedema Stage | 0.144 | 0.223 | 0.416 | 1 | 0.519 | 1.155 |
| Seek Treatment | 0.226 | 0.373 | 0.367 | 1 | 0.545 | 1.254 |
| Stigma | -0.230 | 0.324 | 0.502 | 1 | 0.479 | 0.795 |
| Constant | -2.429 | 1.702 | 2.038 | 1 | 0.153 | 0.088 |

#### Key Associations between Variables

Pearson’s correlation analysis indicated a strong positive correlation between the number of acute attacks and lymphedema severity (r = 0.68, p < 0.001). Furthermore, socioeconomic status was negatively correlated with the presence of nodules (r = -0.42, p < 0.05), indicating that participants from poorer backgrounds were more likely to have nodules.

The study findings revealed that 42.6% (87) of participants reported experiencing stigma, primarily from the community (60.9%) and family (36.8%), highlighting the significant psychosocial burden faced by individuals with lymphedema. This stigma contributes to social isolation and barriers to healthcare access, further exacerbating the challenges of living with the condition. Additionally, participants reported substantial work-related obstacles, with 34.8% (71) indicating that they sometimes faced challenges at work, 28.4% (58) often encountering difficulties, and 21.1% (43) always experiencing obstacles.

## Discussion

### Prevalence of Nodules and Acute Attacks

Our study found that 57.5% of participants reported the presence of nodules, with the majority located in multiple areas of the lower limbs. This finding aligns with previous studies that identified nodules as a hallmark of chronic lymphedema, particularly in advanced disease stages [28]. The high prevalence suggests that nodules play a critical role in disease progression and may serve as an indicator of severe lymphatic dysfunction.

Additionally, 41.2% of participants experienced acute attacks, with an average of 5.89 attacks in the past six months. This finding is consistent with prior research indicating that acute attacks are a frequent and debilitating complication of lymphedema, often triggered by bacterial infections, poor hygiene, or lymphatic obstruction [29]. A study in Ghana also reported similar rates of acute attacks among lymphedema patients, emphasizing the need for improved management strategies [30].

### Association between Nodules and Acute Attacks

A key finding of this study was the strong association between nodules and acute attacks, with individuals with nodules being 3.2 times more likely to experience acute attacks than those without nodules (p = 0.001). This result supports previous findings suggesting that nodules may act as bacterial reservoirs, exacerbate lymphatic obstruction, and contribute to chronic inflammatory responses [31]. Similar associations were observed in studies conducted in India and Brazil, where nodules were linked to increased infection risk and inflammation [32, 33].

From a clinical perspective, this finding underscores the importance of early identification and management of nodules to reduce acute attacks. Effective interventions such as wound care, antibiotic therapy, and surgical removal of nodules could mitigate complications and improve patient outcomes [15]. Future research should explore the long-term impact of these interventions on reducing acute attack frequency.

### Demographic and Clinical Factors

Our analysis indicated that demographic factors such as age, sex, occupation, education, and wealth were not significantly associated with acute attacks. However, males more likely to experience severe lymphedema (26.9%) compared to females (17.8%). This finding aligns with previous studies suggesting that gender differences in disease severity may be influenced by occupational exposure, healthcare-seeking behavior, and biological factors [34]. A study in Ethiopia found that men in labor-intensive jobs were more prone to advanced lymphedema due to prolonged standing and inadequate access to healthcare [27].

### Stigma and Psychosocial Impact

Stigma remains a significant challenge for individuals with lymphedema, with 42.6% of participants in our study reporting experiences of stigma, primarily from the community (60.9%) and family (36.8%). Previous research has documented the profound psychosocial burden of lymphedema, highlighting social isolation, reduced quality of life, and barriers to healthcare access as key consequences [35]. Similar findings have been reported in studies from Uganda and Tanzania, where misconceptions about lymphedema led to discrimination and reluctance to seek treatment [19, 36].

Furthermore, our study found that 34.8% of participants sometimes faced workplace obstacles, while 28.4% often faced challenges and 21.1% always encountered difficulties. This economic burden was particularly pronounced among low-income individuals, such as beggars and daily laborers. Addressing these issues through community education, vocational training, and self-care support programs is crucial to improving patient livelihoods [37].

### Implications for Public Health and Clinical Practice

The findings of this study carry significant implications for both public health and clinical practice. One of the key areas of focus is nodule management, emphasizing the need for early detection and treatment as part of lymphedema care programs. Integrating regular screening, wound care, and surgical interventions where necessary can significantly reduce the risk of acute attacks and improve patient outcomes.

Equally important is patient education, which plays a crucial role in disease management. Raising awareness about proper hygiene, foot care, and early infection treatment can help individuals prevent acute attacks and maintain better health. In addition to medical interventions, addressing the stigma associated with lymphedema is essential. Community-based programs can be instrumental in reducing discrimination, fostering social acceptance, and ultimately improving both psychosocial well-being and access to healthcare services [38].

Beyond health-related concerns, economic support is another vital consideration. Many individuals affected by lymphedema face financial hardships due to limited employment opportunities. Providing vocational training and financial assistance can alleviate this burden, empowering patients to sustain themselves and enhance their overall quality of life [39]. Together, these interventions offer a holistic approach to improving the lives of those affected by lymphedema, bridging the gap between medical care, social inclusion, and economic empowerment.

### Limitations of the Study

Despite the valuable insights offered by this study, certain limitations should be acknowledged. One key constraint is its cross-sectional design, which prevents the establishment of a causal relationship between the presence of nodules and the occurrence of acute attacks. While associations can be identified, determining direct cause-and-effect remains a challenge.

Another limitation lies in the reliance on self-reported data, particularly concerning the frequency of acute attacks. Since participants provided this information based on their recollection, there is a potential for recall bias, which may affect the accuracy and reliability of the findings.

Additionally, the study’s geographic focus presents another consideration. Conducted in Addis Ababa City Administration, the findings may not be fully generalizable to rural areas or other urban settings where healthcare access, environmental factors, and socio-economic conditions differ. As a result, further research in diverse locations is needed to validate and expand upon these findings.

### Future Research Directions

To build on the findings of this study, future research should explore key areas to enhance understanding and improve lymphedema management. Evaluating the effectiveness of different interventions for managing nodules and reducing acute attacks will be crucial [40]. By assessing various treatment approaches, researchers can identify the most impactful methods for improving patient outcomes. Beyond medical interventions, it is also essential to investigate the sociocultural determinants of stigma. Understanding the factors that contribute to discrimination and social exclusion can help develop targeted interventions that foster acceptance and improve healthcare access for individuals with lymphedema.

These findings underscore the urgent need for integrated morbidity management strategies that focus on early detection and treatment of nodules, improved hygiene practices, and patient-centered care. Addressing stigma through community engagement and expanding economic support programs can further improve the quality of life for affected individuals. Future research should explore the long-term impact of nodule management interventions and assess the effectiveness of community-based approaches in reducing stigma and improving healthcare access. Strengthening these efforts can contribute to better disease control and enhanced well-being for individuals living with lymphedema and other neglected tropical diseases.

## Data Availability

All data generated or analyzed during this study are included in this published article.

## Acknowledgements

We sincerely appreciate all study participants for their involvement. Our gratitude also goes to Footwork, the International Podoconiosis Initiative for generously funding the data collection. Additionally, we extend our thanks to the National Podoconiosis Action Network (NaPAN) and the International Orthodox Christian Charity (IOCC) for their support in facilitating the data collection process.

